# RETURN OF VOLUNTARY MOTOR CONTRACTION AFTER COMPLETE SPINAL CORD INJURY: A PILOT HUMAN STUDY ON POLYLAMININ

**DOI:** 10.1101/2024.02.19.24301010

**Authors:** Karla Menezes, Marco Aurelio B. Lima, Denise R. Xerez, João R. L. Menezes, Gustavo S. Holanda, Adriana D. Silva, Eliel S. Leite, Olavo B. Franco, Marcos A. Nascimento, Marco Aurélio Alves Moura de Faria, Cíntia F. Santana, Livia V. Abreu, Renata C. L. Lichtenberger, Maurilio Rosa, Aurélio V. Graça-Souza, Bruno A. Côrtes, Álvaro U. C. Jorge, Ana Cristina Franzoi, Paulo R. Louzada, Arthur S. Ferreira, Rose M. Frajtag, Tatiana Coelho-Sampaio

**Affiliations:** Institute of Biomedical Sciences, Federal University of Rio de Janeiro, Rio de Janeiro, RJ, Brazil; Department of Neurosurgery, Public State Hospital Azevedo Lima, Niteroi, RJ, Brazil; Department of Internal Medicine, Federal University of Rio de Janeiro, Rio de Janeiro, RJ, Brazil; National Cancer Institute, Rio de Janeiro, RJ, Brazil; Institute of Medical Biochemistry, Federal University of Rio de Janeiro, Rio de Janeiro, RJ, Brazil; Department of Neurosurgery, Rio de Janeiro Municipal Hospital Souza Aguiar, Rio de Janeiro, RJ, Brazil; Graduate Program in Rehabilitation Sciences, Augusto Motta University Centre, Rio de Janeiro, RJ, Brazil

**Author notes:** Corresponding author: Institute of Biomedical Sciences, Federal University of Rio de Janeiro, Av. Carlos Chagas Filho, 373, room B1-011, Rio de Janeiro, RJ, Brazil, 21941-902. Department of Neurological Surgery, University of California, San Francisco, CA, USA.

## Abstract

Spinal cord injury (SCI) is still a major challenge for translational medicine. Polylaminin is a stabilized form of laminin, a natural multifunctional protein, previously shown to promote neuroprotection and axonal growth after SCI in rodents. Here we present the results of a confirmatory preclinical study performed in rats and of a first in human trial of polylaminin to treat acute SCI. The latter was an open-label, single-armed academic study designed to include only patients diagnosed with functional complete SCI, because no more than 15% of these patients recover motor function spontaneously. Eight participants received a single intraspinal injection of polylaminin shortly after trauma (average time = 2.3 days). Two died within the first days of causes related to the gravity of the condition. The six patients that survived to reach the one-month follow-up were evaluated using the International Standards for Neurological Classification of Spinal Cord Injury (6/6) and motor and somatosensory evoked potentials (3/6). All the six patients regained voluntary motor control below the level of the lesion, which is an unprecedented recovery. This preliminary study suggests the efficacy of polylaminin to treat acute SCI, revealing its potential as a new therapy to restore motor function to paralyzed people (UTN: U1111-1144-5390).

## INTRODUCTION

An effective treatment for spinal cord injury (SCI) is still an unmet need. The complex and multifaceted pathology of central nervous tissue damage suggests that a successful therapy will require the combination of several therapeutic agents aiming at multiple targets simultaneously [1]. Laminin is an extracellular matrix protein involved in a variety of physiological events in nervous system development and regeneration, including neural cell survival, proliferation, migration, and differentiation [2], which means that laminin can be envisaged as a multitarget agent in itself. Disruption of laminin expression in transgenic mice, when not embryonically lethal, impairs neuronal migration and axonal growth, leads to defects in cortical lamination, disturbs myelination, and affects the integrity of the blood-brain barrier [2-4]. Moreover, laminin expression is a hallmark of successful regeneration in the nervous system [5, 6]. Injured axons in peripheral nerves regenerate inside laminin conduits assembled by Schwann cells, whereas the invertebrate spinal cord rebuilds its axons in contact with laminin [7, 8]. In addition, it has been reported that, when regeneration occurs after experimental SCI, the axons that manage to cross the lesion area do so in contact with laminin [9, 10]. Finally, there are several examples of laminin isoforms promoting neuronal survival [11], modulating neuroinflammation [12, 13] and controlling the biology of the neural stem cell niche [14].

Polylaminin is a stable lab-made polymer of laminin that recapitulates the original structure of laminin assemblies occurring in natural tissues [15]. Polylaminin enhances neurite/axons outgrowth in different neuronal types cultivated *in vitro* [16, 17]. Moreover, an intraparenchymal injection of polylaminin after SCI in rats promoted functional recovery, associated with a decrease in inflammation and tissue loss and to an increase in regenerating axonal fibres [18]. Unlike most preclinical studies that tested potential new therapies using a single type of injury, the efficacy of polylaminin was originally observed using three different types of injury (moderate compression, partial dorsal section, and complete transection) [18]. To further expand the robustness of preclinical data on polylaminin, here we performed a series of 5 independent experiments in rats using a fourth type of injury (mild compression) and confirmed the efficacy of the treatment. The beneficial effect of polylaminin was additionally confirmed by a contracted research organization, using a moderate contusion inflicted by the infinite horizon impactor.

Polylaminin stands as a promising new therapy for SCI in humans as it overcomes the two major barriers in translating preclinical into clinical research, namely, the need for a multitarget therapy and the lack of robustness of animal data [1, 19]. Here we report the results of a pilot study to assess the safety and possible benefits of a single intraparenchymal injection of polylaminin during the acute phase of human SCI.

## RESULTS

### Preclinical study in a rat model of mild balloon compression

To strengthen the preclinical evidence of polylaminin efficacy to treat SCI [18], we performed an additional animal study using a mild balloon compression injury [20]. Importantly, because in previous studies the efficacy of polylaminin had been tested by comparing the locomotor performance of treated animals with that of animals receiving injection of vehicle, we judged crucial to confirm that the benefits of the drug would stand when the comparison were made against non-injected controls, which would more adequately correspond to non-treated SCI patients. Accordingly, here we used a total of 106 rats, among which 51 received a single intraparenchymal injection of polylaminin and 55 did not receive any injection. Functional outcomes were assessed by evaluating open field locomotion (BBB scores) [21] across 8 weeks (Fig. 1A). At 24 hours and after 1 week there was no difference between the two groups. However, after 2 weeks until the end of the experiment animals receiving polylaminin presented significantly higher scores (p < 0.01). Interestingly, at the 8^th^ week after lesion, rats treated with polylaminin reached an average BBB score of 19.6±0.3, which is close to the score of 20, that corresponds to full gait capacity with only mild instability of the trunk, while control animals stabilized at 18.0±0.4. These results demonstrate that rats treated with polylaminin after mild SCI show better motor performance than rats not receiving any treatment.

**Figure 1.**
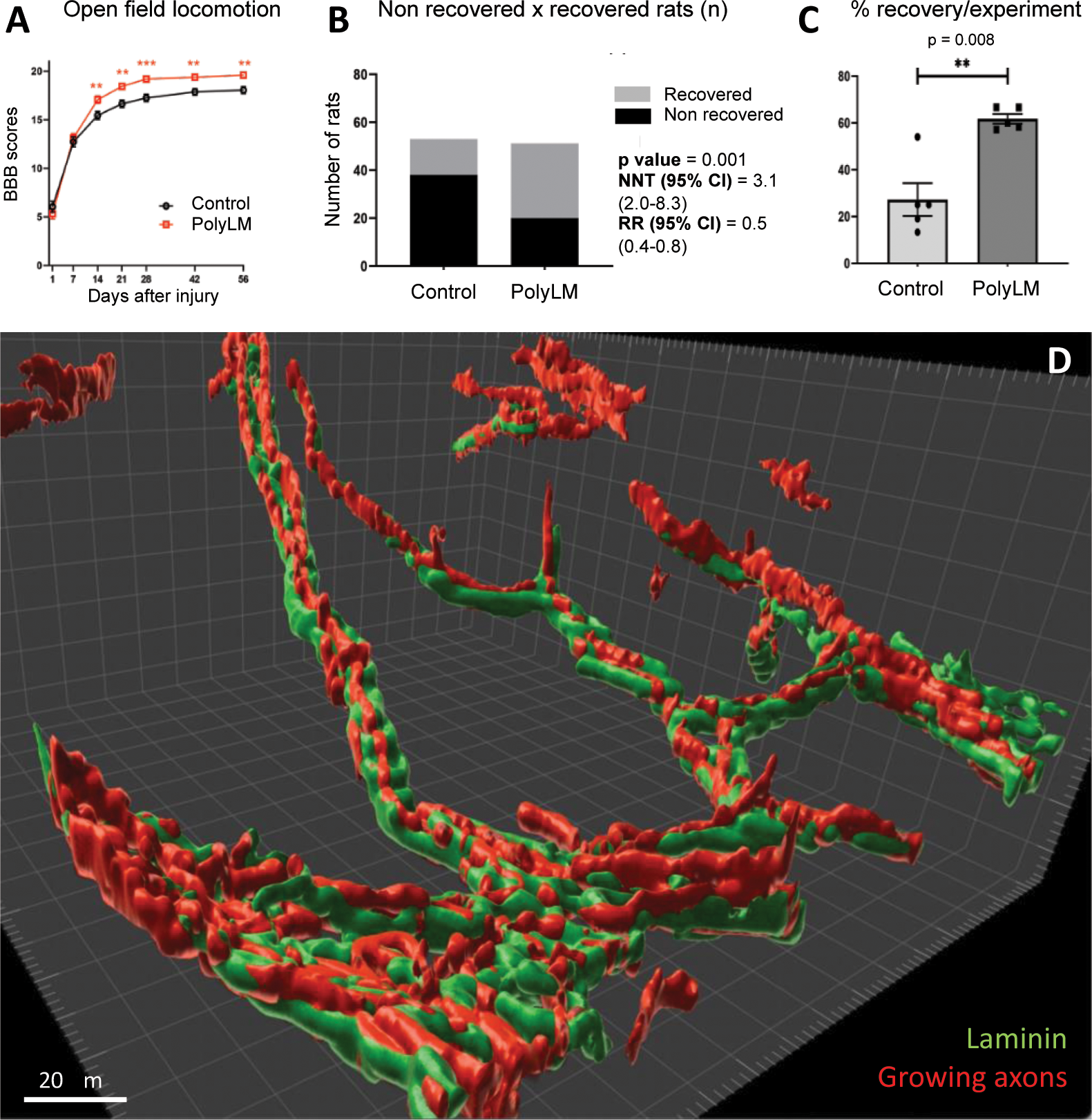
Polylaminin improves the outcome of experimental SCI in rats. (**A**) Open field locomotion was evaluated in rats submitted to a mild compressive lesion and treated with polylaminin (PolyLM; red squares) or not receiving any injection (black circles). Values of BBB scores correspond to means±SEM. (**B**) The numbers of animals recovered and non-recovered are compared between the control and polyLM-treated groups. (**C**) The percentages of recovered and non-recovered rats are presented for each experimental round (N values of 12/12, 14/11, 12/15, 10/13 and 3/4 for polylaminin-treated/non-treated). (**D**) Three-dimensional reconstruction (Imaris software) of axons growing through the lesion site in a treated animal. Axons (red) were immunostained with a marker of plastic neurons (TUJ-1 antibody). Young axonal fibres crossing the injury site are in contact with laminin-positive conduit-like structures that seem to pave their way across the lesion gap (green). **, p < 0.01 and ***, p < 0.001.

Next, we reassessed the same data to compare the total numbers of animals recovered (scores 20 and 21) and non-recovered between the two groups (Fig. 1B). We found that the proportions of non-recovered animals in polylaminin-treated, and control groups were 0.39 and 0.72, respectively, providing a Risk Ratio (RR) of 0.5 with 95% confidence interval of 0.4 to 0.8. The Number Needed to Treat (NNT) calculated was 3.1 with a 95% confidence interval between 2.0 and 8.3.

As the preclinical study (total N value of 106) was performed in 5 separated rounds (N values of 24, 25, 27, 23 and 7 for each round), we additionally compared the percentages of recovered and non-recovered rats across individual experiments. As seen in Figure 1C, the mean ± SEM of the recovered rats shifted from 27.3 ± 16% to 61.8 ± 5% when animals were treated with polylaminin. Taken together, data collected from functional analyses in the preclinical study confirm the potential benefit of polylaminin for SCI.

A qualitative morphological analysis was also performed. Representative images of spinal cords in each experimental group are shown in Fig. S1. Panel D in Fig. 1 depicts the strict correlation between axonal regeneration across the lesion epicentre and the presence of laminin in a rat treated with polylaminin. Axons growing across the lesion, do it on gutter-shaped laminin conduits. While we cannot determine whether laminin making these conduits correspond to the laminin injected or to laminin produced endogenously by the animal (the polyclonal antibody does not distinguish between the two), we can clearly appreciate the pivotal role of laminin in supporting axonal regrowth across the lesion site. The confirmatory preclinical data reported here support polylaminin as a valuable therapy to be tested in a human study.

### Demography of the human trial

Eight participants were recruited between December 2016 and January 2021 and treated within the following 6 days, 2.3 days being the mean time for polylaminin injection. Demographic analysis of the participants is shown in Table 1. They were 5 males and 3 females (62,5% male), aged 23 to 69 (average 39,1). Causes of the injury were fall (4), motor vehicle accident (2) and transfixing gunshot (2), at neurological levels C5 to T10; 4 were tetraplegics and 4 were paraplegics.

**Table 1.** Demography of the participants.

| *Participant* | *Age* | *Sex* | *Cause* | *Neurological level* | *Date of injury* | *Time for treatment* |
| --- | --- | --- | --- | --- | --- | --- |
| 1 | 30-35 | F | Fall | C7 | Dec 2016 | 1 day |
| 2 | 20-25 | M | Motor vehicle accident | C7 | Apr 2018 | 1 day |
| 3 | 30-35 | M | Transfixing gunshot | C4 | Sep 2018 | 2 days |
| 4 | 55-60 | F | Fall | T2 | Sep 2018 | 3 days |
| 5 | 30-35 | M | Transfixing gunshot | T10 | Oct 2018 | 1 day |
| 6 | 30-35 | M | Motor vehicle accident | C5 | Oct 2019 | 4 days |
| 7 | 40-45 | F | Fall | T10 | Sep 2020 | 6 days |
| 8 | 65-70 | M | Fall after electrical shock | T4 | May 2021 | Less than 24h |

Seven of the 8 participants received early decompression/stabilization surgeries. Only participant 3 had no surgery indication and the treatment was delivered percutaneously. Participant 6 also received a percutaneous injection because study enrolment occurred when the surgery had already been carried out. Figure S2 shows representative frames of the initial image examinations of the participants.

### Medical complications in the human trial

All adverse events occurring from the time of inclusion until the end of the study were readily treated and their occurrence, recorded in the study data basis. After completion of the trial, adverse events were analysed by an external physician (GSH) according to the classification proposed by the NACTN SCI Registry [22]. The events are listed and classified in Table 2.

**Table 2.** Medical complications classified by organ/system and severity. Medical complications during hospitalization were monitored in the 8 patients and classified by organ/system and severity. ^1^Refers to one case of lower limb oedema (moderate) and one case of poorly positioned arthrodesis equipment (mild).

| Type of complication | Total no. of complications |  |  |  | No. of patients |
| --- | --- | --- | --- | --- | --- |
|  | *severe* | *moderate* | *mild* | total |  |
| Infectious | 8 | 5 | 1 | 14 | 7 |
| GI/GU | 4 | 3 | 8 | 15 | 7 |
| Neuropsychiatric | 3 | 5 | 6 | 14 | 7 |
| Skin | 1 | 3 | 1 | 4 | 4 |
| Cardiovascular | 8 | 1 | 1 | 10 | 3 |
| Pulmonary | 6 | 3 | 0 | 9 | 3 |
| Haematological | 2 | 0 | 0 | 2 | 2 |
| Others<sup>1</sup> | 0 | 1 | 1 | 2 | 2 |
| Total | 32 | 21 | 18 | 71 |  |
| % Total | 45 | 30 | 25 | 100 |  |

During hospitalization, all participants experienced some adverse event. Seven participants presented at least one infectious event within one month after injury. Eight out of a total of 14 infection complications were considered severe. The second and third most common occurrences were gastrointestinal/genitourinary (GI/GU) and neuropsychiatric complications, also affecting 7 participants but involving less severe medical events. Four participants presented skin complications, mostly pressure sores. Three out of 7 patients had cardiovascular and pulmonary complications, while haematological adverse events were observed in 2 participants.

Severe complications are described in detail in Table S1. When only severe events were considered, infections remained the leading complication, bearing an incidence of 63% (5 out of 8 participants). Pulmonary and GI/GU complications had incidences of 38%, meaning that 3 participants out of 8 had at least one complication in these categories.

Cardiovascular, neurological/psychiatric, and haematological adverse effects occurred in 25% of patients (2 out of 8), and severe skin ulcers during hospitalization, in 17% (1 out of 8). A list of all complications is provided in Table S2. None of the adverse events could be ascribed to the treatment (see Discussion).

Among the 8 participants, two evolved to death during hospitalization. Participant 1 presented signs of broncho aspiration from admission and developed severe pneumonia within the first days of hospitalization. Death occurred on the fifth day after treatment. Participant 3 had a sudden cardiovascular event on the 20th day of the follow-up, coursing with pericardial effusion and cardiac tamponade. It was hypothesized that effusion was caused by a fragment of the projectile that had hit the pericardium at the accident. Participant 8 was discharged after 3 weeks of hospitalization, went through the 1-month follow-up at home (during the pandemic), but was hospitalized again due to an infected skin ulcer. He died of sepsis two months after inclusion. The Brazilian National Commission of Ethics in Medical Research (CONEP) concluded that deaths were not related to the treatment.

### Monitorization of hepatic and renal markers of toxicity in the human trial

Blood tests were performed before treatment and compared with tests carried out 2 and 10 days after treatment to monitor markers of renal and hepatic toxicities (Table S3).

Creatinine and urea were evaluated in all participants (8) and the sole deviations were mild elevations of less than 1.2-fold of the reference values observed in creatinine for participant 8 (elevation detected in the 3 assessments, including the one before treatment) and in urea for participant 6 (at 2 days). For alanine and aspartate aminotransferases (ALT and AST) most of the deviations detected were considered mild. The only two moderate elevations were already present before treatment and disappeared on the 10th-day evaluation (participants 2 and 8). No severe elevations of liver enzymes were found. In addition, no important deviations were detected in blood counts, except for a mild anaemia within the first days after surgery (Table S4).

### Neurological evaluation in the human trial

Efficacy of polylaminin was assessed by neurological examinations using the ASIA Impairment Scale (AIS). The use of a standardized scale is what validates the design of single-arm trials, in which the data collected from a group of individuals, all receiving the treatment, can be compared with a universal baseline established in previous longitudinal studies that used the same scale to assess the spontaneous recovery of patients not receiving any treatment [23, 24]. It is well established that no more than 15% of individuals bearing complete injuries (AIS A) will spontaneously recover motor function (AIS C, D or E) after 1 year [23-25]. Table 2 shows the evolution of AIS grades along the 12 months follow-up.

**Table 2.**
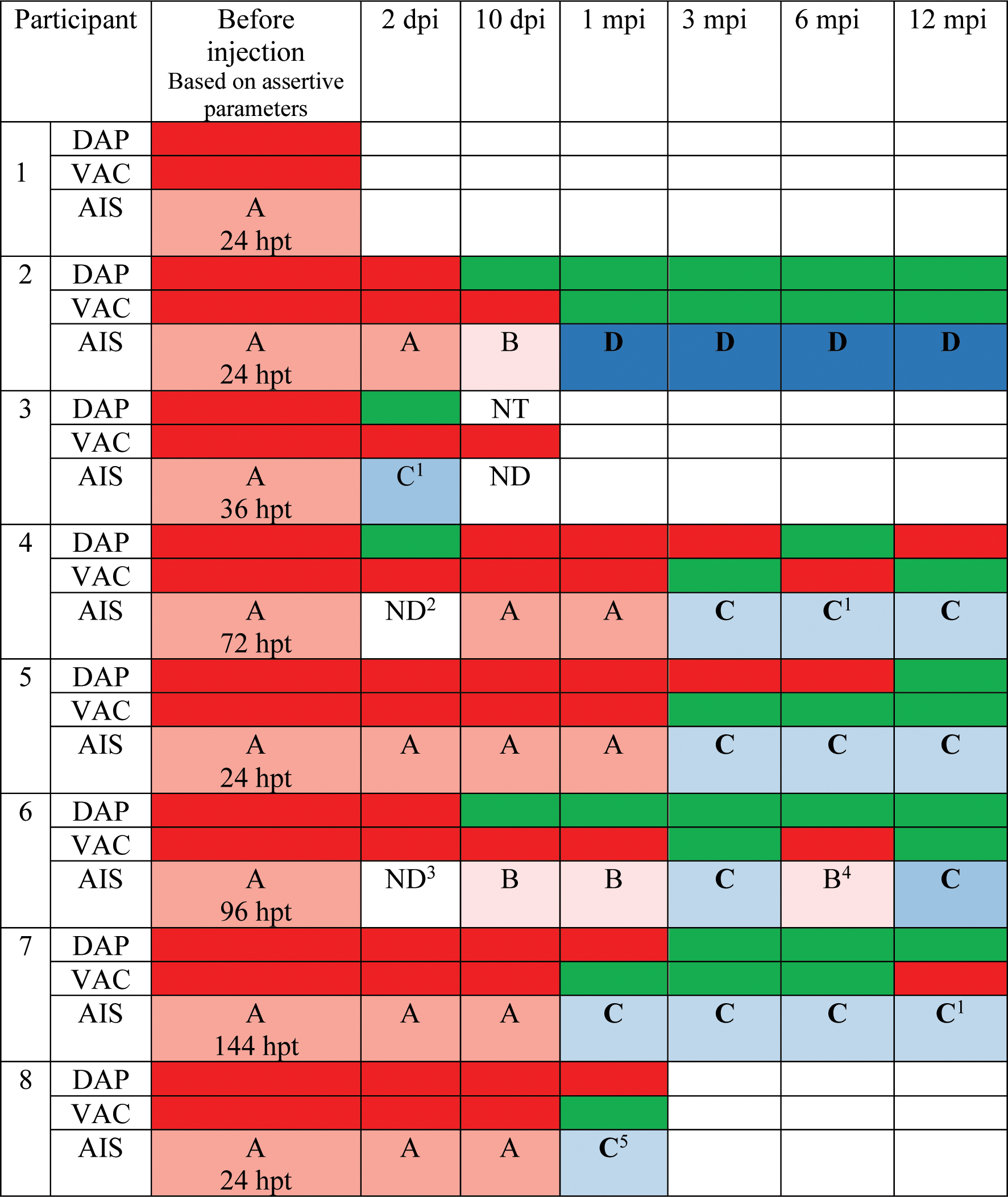
Neurological evaluation of the 8 participants in the human trial. ^1^DAP preserved and some sparing of motor function more than three levels below the ipsilateral motor level; ^2^PP and LT not assessable at the cervical region due to collar immobilization; ^3^Patient sedated; ^4^No VAC was detected, but patient reported pain due to an anal fissure; ^5^DAP preserved and some sparing of motor function more than three levels below the ipsilateral motor level, and contraction of non-key muscles in L4. Participants 1 and 3 died during hospitalization and participant 8, after discharge. Abbreviations: dpi and mpi apply to days and months post-injection, respectively. ND means “not determined,” because examination results were not sufficient to determine. NT means “not testable,” used here because the patient was not able to clearly discern between yes and no. Hpt refers to hours post-trauma. DAP and VAC stand for the sensation of deep anal pressure and voluntary anal contraction, respectively, and were represented by the red or green colours when absent or present, respectively. For the AIS grades we used the following colour code for emphasis: dark pink for AIS A, light pink for AIS B; light blue for AIS C, and dark blue for AIS D.

Three participants (2, 7, 8) regained motor control. i.e., converted to AIS C or D, at the 1-month examination. Other three (4, 5, 6) converted to grade C after 3 months. In the present study we observed that, in contrast to the expected baseline conversion of 15%, 75% (6/8) of the participants regained voluntary motor control after polylaminin treatment. If we consider only those participants that reached discharge, the proportion increases to 100% (6/6). The fact that patients treated with polylaminin recovered motor function above the baseline of spontaneous recovery suggests its efficacy to treat spinal cord injury. Motor and sensory scores of the 5 participants completing the follow-up are shown in Fig. 2.

**Figure 2.**
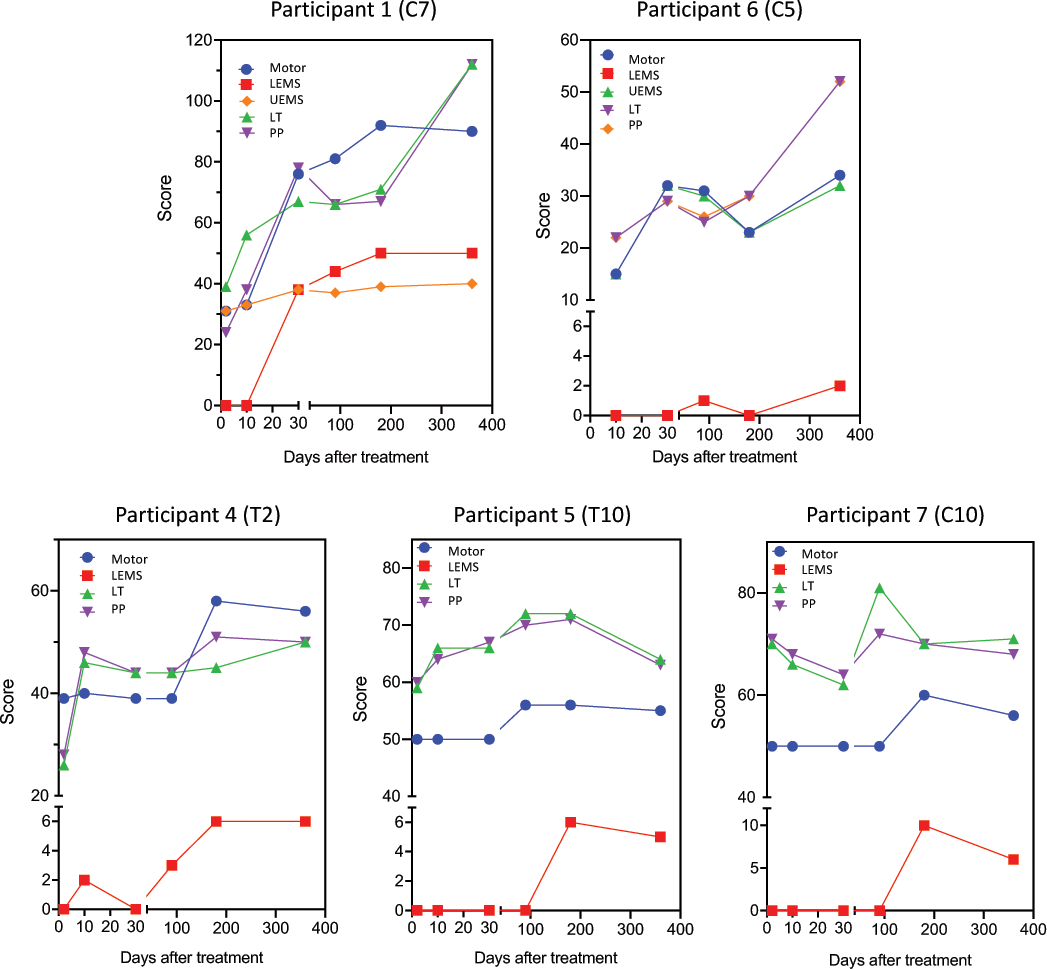
Motor and sensitive scores. Total motor scores (Motor), lower extremity motor scores (LEMS), upper extremity motor scores (UEMS), light touch (LT) and pin prick sensitivities (PP) were registered during the follow up visits at 2, 10, 30, 90, 180 and 360 days. Graphs are grouped as cervical (upper panels) and thoracic (lower panels). Note that we used different y axes to highlight the observed changes.

Recoveries of both sensory and motor scores were more pronounced in participants with cervical lesions. Participant 2 recovered normal sensitivity across all dermatomes, regained independent control of bladder and bowel function, and attained full walking capacity. At the final evaluation, participant 6 had improved his neurological level from C5 to C7. Besides VAC and DAP, he recovered sensitivity bilaterally at S3, S4-5, became able to localize tactile stimulus on the muscles of both legs, and gained voluntary contraction at C8, T1, L2, and S1 on the left. Along the follow-up, he showed voluntary contraction in alternated key and non-key muscles. Among the paraplegic participants, we found little recovery of sensory function other than DAP identification (Fig. 2). Lower extremity motor scores improved from the third month (participant 4) or from the sixth month of follow-up (participants 5 and 7). Gains of 6 to 10 points were documented for these three patients, although a tendency to stabilization or reduction was observed in the 12-month evaluation. This was a relatively small but consistent gain, considering that the progression of LEMS in paraplegic patients is rare [23].

### Imaging of SCI patients

The 5 participants who completed the 12-month follow-up had their initial and final imaging exams compared. Participant 2 developed a large cystic cavity within the spinal cord that occupied most of its cross-section. (Fig. 3A). Participant 4 also developed a cavity at the injury site (Fig. 3B). Participant 6 at 12 months presented only a narrow strip of normal signal remaining at the centre of the spinal cord (Fig. 3C). Participant 5 did not undergo MRI due to the presence of metallic fragments of the projectile. Comparison of CT images did not reveal any remarkable change during the follow-up period. Participant 7 had only the final MRI performed because there was no MRI machine in the hospitalization centre, and it was not possible to transfer the patient for the exam during the peak of the pandemic. Analysis of the final MRI image also indicated the presence of a large area of hypersignal at the lesion site. Overall, image analyses suggest that polylaminin treatment did not lead to any image-detectable deviation of the cavitation process expected to occur after SCI [26, 27].

**Figure 3.**
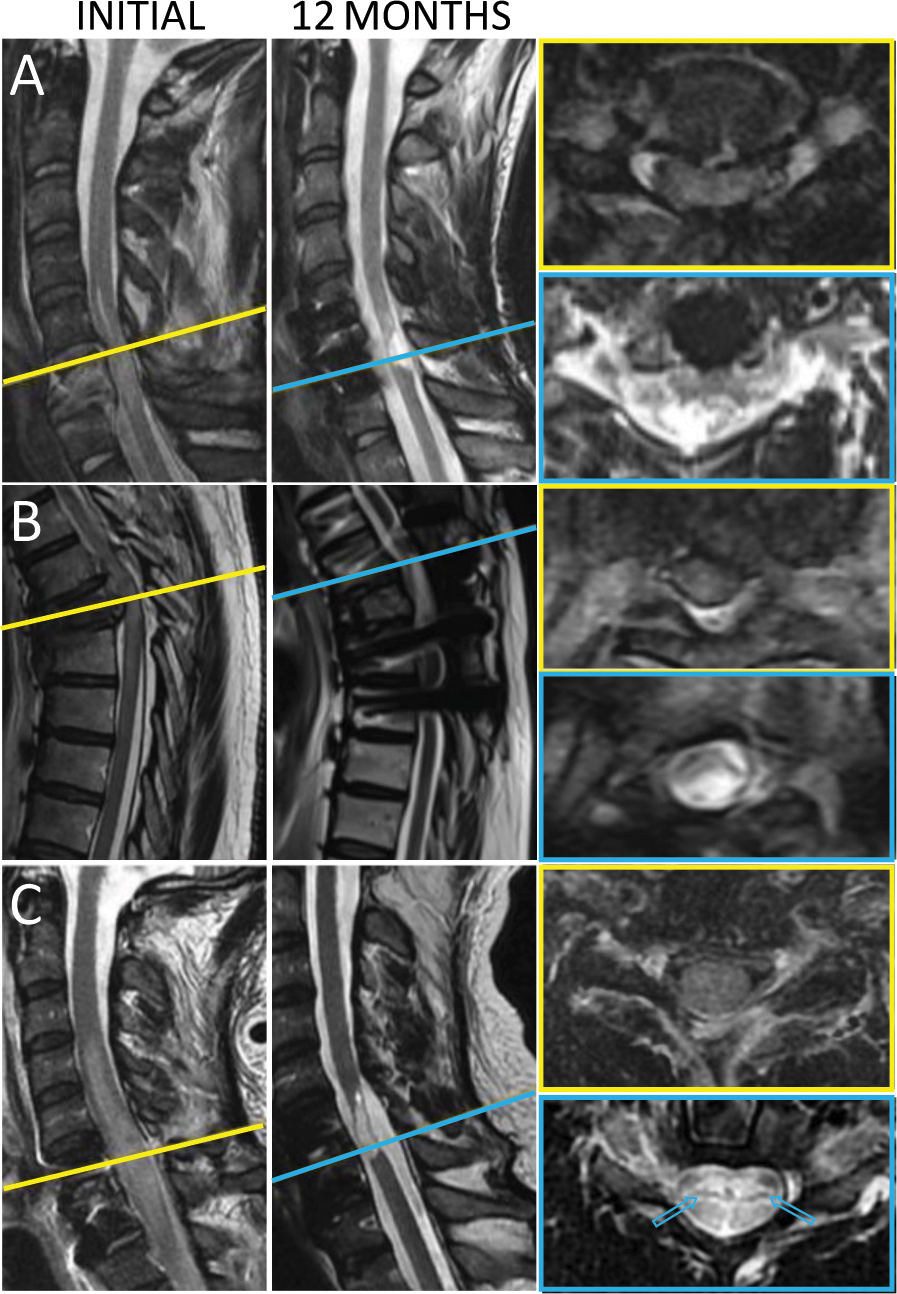
Initial and final MRI frames. **A)** T2 sagittal and axial sequences of participant 2 performed before and 12 months after polylaminin injection. The initial image shows the fractured C6 vertebra and the oedema spreading across the whole section of the spinal cord. At 12 months an oval-shaped area of hypersignal, compatible with the presence of liquid, occupied the lesion site. **B**) T2 sagittal and axial sequences of participant 4, before and 12 months after polylaminin injection. The final MRI of participant 4 was affected by the artifacts generated by the prothesis material. Nevertheless, the image is compatible with the formation of a cystic cavity at the injury site. **C**) MRI frames of participant 6 are shown in T2 sagittal and axial sequences before polylaminin injection and at the end of the 12-months follow-up. In the initial image an extensive area of oedema occupies the whole section of the spinal cord, leading to complete interruption of liquor flux. The final image shows intense hypersignal along the lesion area, contiguous to a narrow strip of preserved tissue (arrows in axial view).

### Electrophysiological assessment of SCI patients

Electrophysiological evaluations were performed either by surface electromyography or evoked potentials. Participants 2, 4 and 5 were submitted to surface electromyography (Supplemental Material). Participant 2 showed a progress compatible with that observed in the neurological evaluation. Participants 4 and 5, presented no alterations between the initial and the 12-month exams, meaning that the electromyography was not sufficiently sensitive to detect the voluntary contractions observed in the final neurological examinations of these participants (see LEMS values in Fig. 2).

Participants 2, 6, and 7 received motor and somatosensory evoked potentials examinations (MEP and SSEP, respectively). In participant 2 initial SSEP patency was found only in the upper limb on the right, while MEP was normal only down to C7 (Fig. 4A). No evoked potentials were observed in lower limbs. MEP at the end of the follow-up showed the return of potentials, with or without facilitation, in hand muscles bilaterally (little finger abductor; T1), although with delayed latency. Motor potentials with delayed latencies also returned at tibial and adductor hallucis muscles (L4, L5 and S1) even without facilitation (Fig. 4A). Final SSEP was evaluated at the median and tibial nerves and was found abnormal but present bilaterally in both nerves.

**Figure 4.**
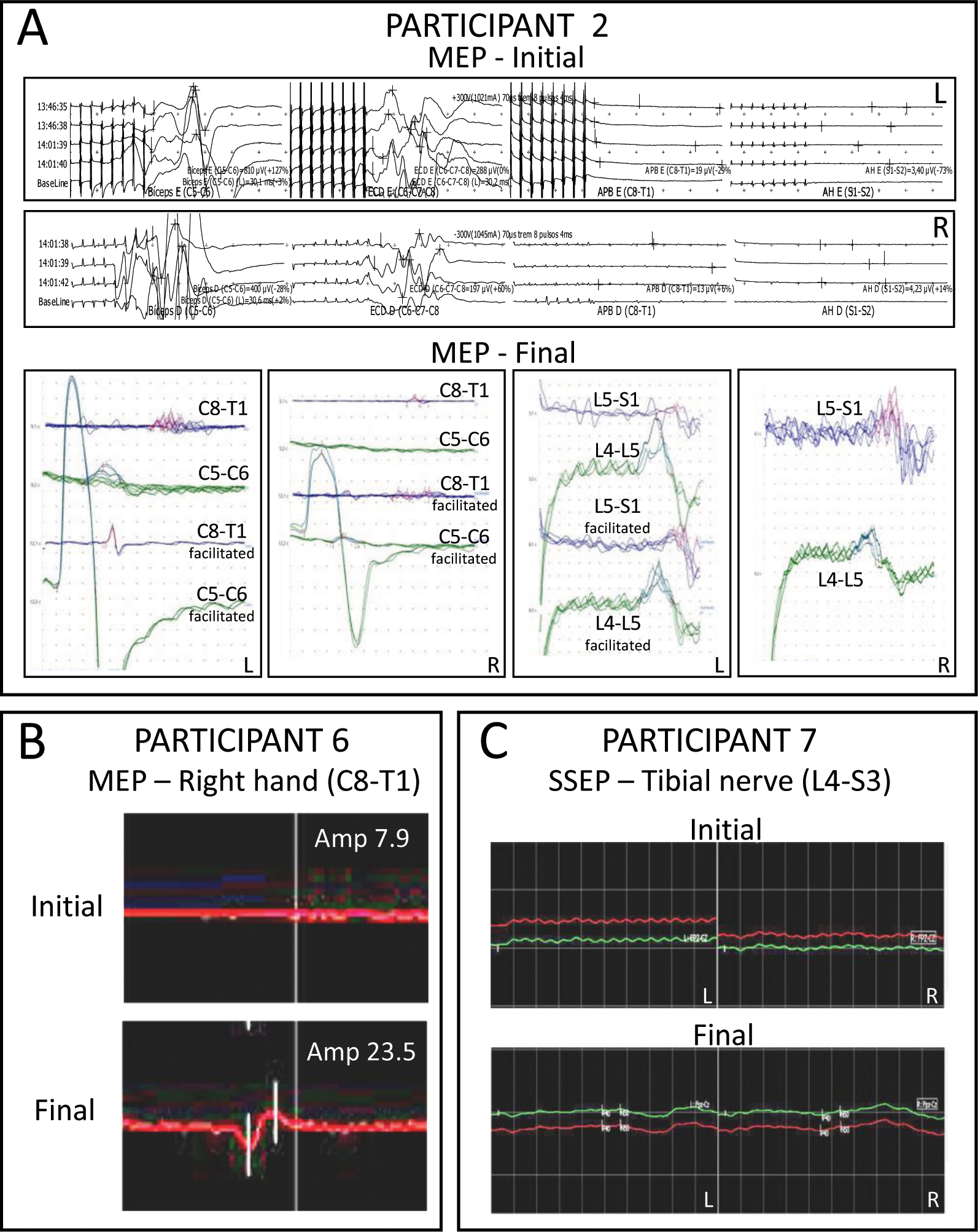
Electrophysiological monitoring. Frames extracted from the original evoked potential recordings of participants 2, 6 and 7. **A**) MEPs of participant 2 were measured on Neurosoft equipment, using transcranial electrical stimulation at points C3 and C4 (10/20 system) and captured at the following muscles: biceps, common finger extensors, pollicis brevis abductors, and hallucis abductor (left to right). Final MEP was recorded using transcranial magnetic stimulation in Neurosoft Neuro-MEP-Micro equipment. Capture was performed at the deltoid, little finger abductor, anterior tibialis and hallucis abductor muscles. **B**) Motor evoked potentials of participant 6 were recorded with a Medtronic NIM Eclipse 32 channels equipment, using transcranial electrical stimulation at points C3 and C4 (10/20 system) and capture at the distal muscles of the upper limbs (intrinsic hand muscles). **C**) Somatosensory evoked potentials of participant 7 were assessed with stimulation of tibial nerves at ankles (P40/N50) bilaterally and capture in the skull at points CZ’-FPZ and FPZ-C3; FPZ-C4 (10/20 system). L and R stand for left and right, respectively.

The initial SSEP of participant 6 detected normal conduction in median nerves (C6) and reduced amplitudes in the tibial nerve (L4-S3). MEP was present and symmetrical down to C6-C7 and absent at C8-T1 and in lower limbs. At the end of the follow-up, SSEP remained unaltered, but MEP registered a previously absent potential at C8-T1 on the right hand (Fig. 4B).

Participant 7 bearing a T10 injury presented normal SSEP and MEP in upper limbs at all examinations. The initial SSEP revealed an unequivocal loss of patency in lower limbs bilaterally. On the other hand, the final assessment indicated the presence of low amplitude potentials identified by the P40/N50 markers at tibial nerves (Fig. 4C). MEP remained unaltered across the 12 months.

## DISCUSSION

Here we report that the acute treatment with polylaminin leads to the return of voluntary motor contraction in 6 of 8 patients diagnosed with functional complete SCI (AIS A). The conversion from AIS A to C was observed in 5 of 8 participants in this pilot trial, while conversion from AIS A to D was observed in 1 of 8 participants. Since the two subjects that did not reacquire motor function were those who died during hospitalization, we can interpret that 100% of the patients (6) who survived to reach at least the 1-month follow-up regained motor control.

Previous clinical trials with larger numbers of patients have assessed the efficacy of new therapies for the acute phase of SCI. Like ours, most of them were uncontrolled, single-armed studies, in which efficacy was to be established in comparison with the baseline of SCI natural history reported for untreated patients (between 0 and 15% across different studies) [23, 25, 28]. Acute treatments with the cytokine, granulocyte-colony stimulating factor [29], with the sodium-channel blocker, riluzole [30] or with the anti-NOGO antibody, ATI355 [31], resulted in, respectively, 0, 16 and 6% of AIS A participants converting to C or D, which did not correspond to a progression above the baseline observed in untreated patients. On the other hand, injection of autologous activated macrophages [32] or treatment with the Rho-A inhibitor, Cethrin [33], led to noteworthy conversions from complete to motor incomplete of 38% and 30%, respectively.

Polylaminin is derived from a natural extracellular protein ubiquitously distributed across the body. Accordingly, the adverse events encountered in the study subjects were qualitatively like those reported in the literature for the same condition. Spinal cord injury patients undergo dysautonomia and alterations in cardiopulmonary function, which extensively affects normal physiology [34]. We found that all participants (8/8) presented at least one medical complication during hospitalization. Although this is a high prevalence, it is comparable to the 84 and 63% previously reported for AIS A patients [22, 35].

We had 3 deaths among the 8 participants, caused by pneumonia, pericardial effusion, and sepsis, whereas the first two occurred during hospitalization and the third, one month after discharge. Hemodynamic alterations due to the loss of vasoconstrictor tone, as well as impairment of the diaphragmatic and intercostal muscles, predispose SCI patients to cardiovascular dysfunction and to pneumonia, the latter being the leading cause of mortality in these patients [34]. Nevertheless, pneumonia and sepsis (participants 1 and 8, respectively) could in principle be related to an eventual sustained anti-inflammatory state provoked by the treatment, as polylaminin has been shown to display anti-inflammatory properties [18]. At this point, we cannot completely rule out this possibility. On the other hand, examination of medical records of the 8 participants did not reveal signs of leukopenia or reduction of C-reactive protein below the expected levels during the post-operatory window (Table S4), which weakens this hypothesis. In the three cases (participants 1, 3, and 8), the series of medical events leading to death were evaluated and the outcomes were considered related to their clinical conditions. A recent study carried out in emergency centres in Japan observed that 43.8% of cervical SCI patients classified as AIS A progressed to death [36]. A Brazilian retrospective study reported a mortality of 46% for patients with cervical injuries [37]. Another study carried out between 1996 and 2011 in public hospitals in Rio de Janeiro, the same setting of the present study, revealed that the mortality of SCI patients (all AIS grades and neurological levels pooled together) varied from less than 5% to more than 40%, depending on the year considered and on whether the hospitalization centre was managed by the state or the municipality [38].

Our finding that a single-dose therapy was effective in promoting functional recovery after SCI indicates that a short-term contact with polylaminin is sufficient to impel the damaged tissue into a pro-regenerative mode. In this regard it has been previously shown that a single encounter with laminin dramatically accelerates axonal growth of sensory neurons *in vitro* [39]. An alternative explanation is that the injected protein, particularly in the polymerized form, remains active in the spinal cord for sufficient time to support elongation of the fibres sprouting after lesion. Another intriguing point is that the functional recovery observed in the present study occurred without parallel signs of robust tissue regeneration that could be detected by image analyses. This does not rule out motor and sensory recovery due to axonal regeneration, as it is possible that few regenerating axons could be enough to input pattern rhythm over intrinsic medullary pattern generator centres [40, 41]. It has been shown that a conserved hierarchical integration of local central pattern generators can harmonize segmental networks for ambulation, movement, and autonomous control [42]. One can conceive that few descending serotonergic or adrenergic pathways could impose descending rhythmic control, much like electric stimulation applied epidurally to distal segments of SCI can efficiently induce rhythmic movement in paralyzed humans [43, 44].

The preliminary data reported in the present study suggest that polylaminin can work as a biomimetic replacement for the axonal-growth permissive laminin missing in mammalian CNS after lesion. In our view, the remarkable functional improvement experienced by the patients of this pilot study warrants the design of larger clinical trials to test polylaminin as a therapy for human SCI.

## MATERIALS AND METHODS

### Preclinical study in rats

Preclinical data were obtained using a rat model of compressive lesion [20]. The protocol was revised and approved by the Animal Welfare Committee of the Federal University of Rio de Janeiro/Centre of Health Sciences (A10/21-042-21). Wistar female rats weighted 200-250g, bred at the animal facility of University of São Paulo (FMUSP; São Paulo, SP, Brazil), were kept in 12h light/dark cycles, with free access to food and water in microisolator cages. Animals were submitted to laminectomy at T7 vertebra and had a Fogarty 2F catheter introduced along the vertebral canal until the balloon reached the T8/9 level. The balloon was inflated with 10 μl of water for 5 min to produce a mild compression. After removal of the catheter, 10 μl of polylaminin (100 μg/ml; 4 to 5 μg/kg) were injected into the cord at T7 placing the needle at an angle of 45° to the horizontal aiming at the preferential diffusion of the liquid to the caudal direction to reach T8/9 (approximately 1 cm). The control group received laminectomy and lesion only (no injection). In order to purposely introduce experimental variability to the study, we divided the experiment in 5 rounds, performed at least one month apart from each other, using animals from different broodings, and ground transported separately between the bred facility and our laboratory. In addition, the experimenters carrying out each procedure (anaesthesia, laminectomy, lesion, injection, suture and post operative care) were alternated in each round. Open field locomotion (BBB scores) was evaluated by two examiners at 24 hr and at 1-, 2-, 3-, 4-, 6- and 8-weeks post injury, with the examiners blind to the experimental group of the animal. BBB scores were used either to calculate means and standard errors for the complete experiment (aggregated values in the 5 rounds; Fig. 1A) or to reckon the number of animals reaching full locomotor recovery, here defined as the sum of those graded BBB scores of 20 and 21. The difference between these two scores relates to subtle changes in trunk stability, which do not impact in gait quality, meaning that rats receiving a score of 20 have full deambulatory capacity [21]. The total numbers of recovered *vs.* non-recovered rats were then used to assess Risk Ratio (RR) and the Number Needed to Treat (NNT) (Fig. 1B). Additionally, we calculated the percentages of recovered and non-recovered animals for each of the 5 experimental rounds, which allowed us to evaluate the efficacy of polylaminin across the individual experiments, among which we had introduced experimental variability (Fig. 1C). As for the statistics, BBB scores were compared using the student’s t test; percentages of totally recuperated, by two-tailed Mann-Whitney test, and the number of animals recovered and non-recovered were compared using the Fisher’s exact test.

At the end of the experiment, the spinal cords of the 10^th^, 20^th^ and 30^th^ animals in each group were sampled and prepared for morphological analyses. Rats were transcardially perfused with paraformaldehyde 4% and the vertebral columns were removed, post-fixed for 4 days and sectioned in a vibratome. Fifty micrometres’ sections were immunostained to reveal young axon fibres, using the anti-beta tubulin III antibody TUJ-1 (R&D Systems; MAB1195; 1:200), and laminin, using a polyclonal anti-laminin antibody (Millipore-Sigma; L9393; 1:50). Observation was done in a Zeiss Elyra PS-1 microscope and z-stacks of confocal images were collected at the lesion epicentre.

### Design of the Clinical trial

This academic clinical trial was approved by the National Commission of Ethics in Medical Research (CONEP), Brazil, CAAE 3.474.884, and retrospectively registered in the Brazilian National Clinical Trials Registry under the Universal Trial Number (UTN), U1111-1144-5390. The present study was supported by academic research grants and not sponsored by the industry.

This was an investigator-initiated, prospective, multi-site, single arm, open-label, pilot trial designed to assess the safety and efficacy of an acute intraspinal injection of polylaminin in individuals diagnosed with functional complete SCI (AIS A). Safety was evaluated by clinical examination, laboratory tests, and monitorization of medical complications. Efficacy was defined as a qualitative elevation from the expected baseline for natural history of complete SCI (AIS A) established in previous studies [23, 25, 28]. These studies demonstrated that the percentage of AIS A patients that spontaneously regained volitional muscle contraction (AIS C, D or E) varied between 0 and 15%, meaning that the vast majority remained AIS A or reacquired only some sensitivity without motor function (AIS B).

The participant hospitals are provided in Table S5. Inclusion and exclusion criteria are listed in Table S6. Follow-up visits were carried out on days 2±1, 10±2, 30±4, 90±20, 180±30, and 360±30 after injection. The flow chart of the study is shown in Fig. 5.

**Figure 5.**
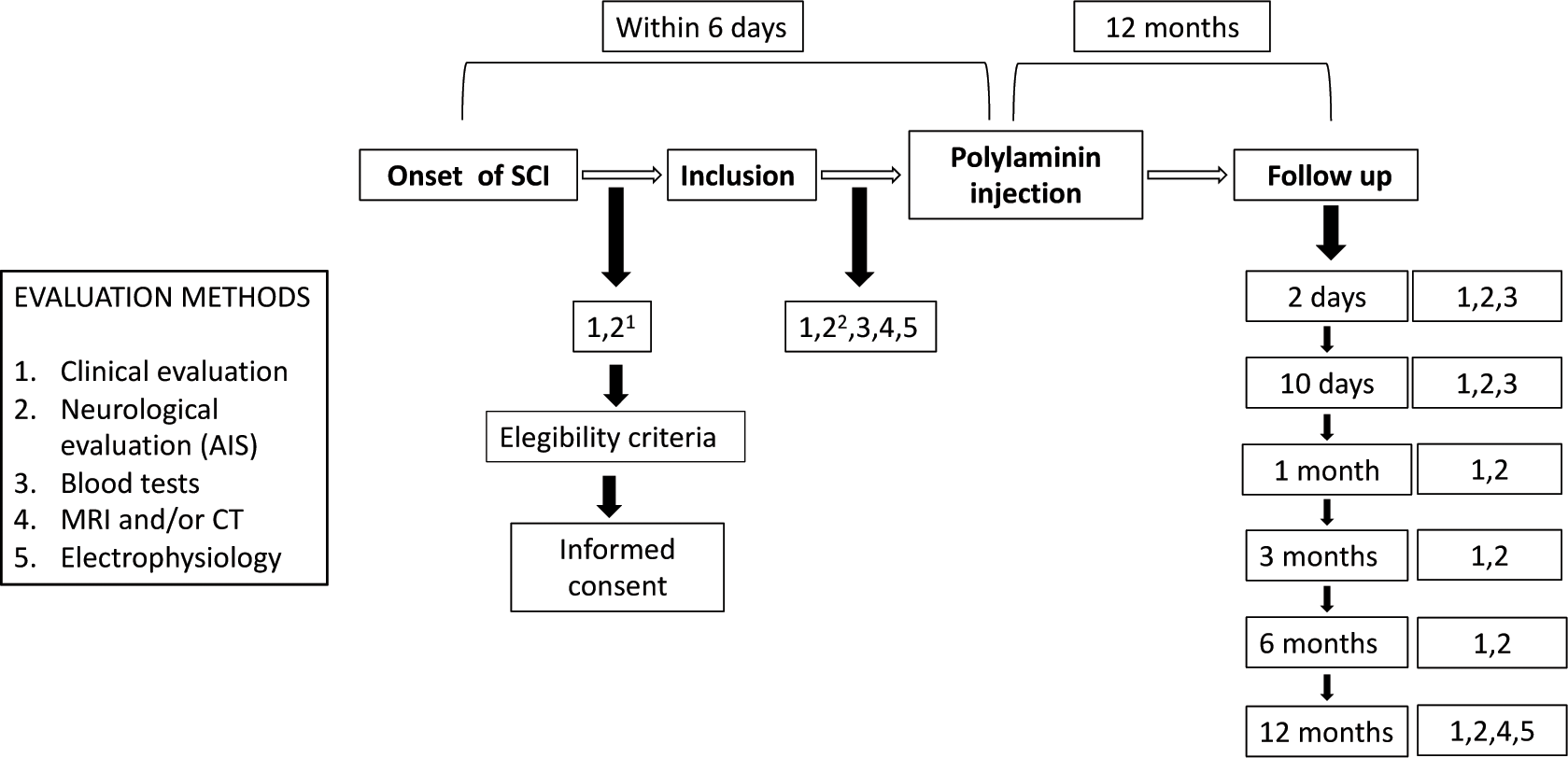
Flow chart of the study. ^1^AIS was evaluated by the on-call physician to define adhesion to inclusion criteria. ^2^AIS was confirmed immediately before treatment by one of the study physicians (see details in the next section below).

After discharge, the study offered 200 h of neurofunctional physiotherapy, encompassing techniques of neuromyofascial mobilization, proprioceptive neuromuscular facilitation/PNF-Kabat method, Bobath concept, Pilates techniques, intensive and continuous rehabilitation, and orthopaedic manual therapy. As it is well established that rehabilitation alone is not able to restore motor function to AIS A patients [45], participants were left free to engage in any additional rehabilitation program of his/her choice.

### Patients

Informed consents were obtained from all participants after the nature and possible consequences of the study were explained. The present study included only patients graded AIS A because these patients are not expected to recover motor function spontaneously. Since the eventual positive outcome of the study was defined as the progression from AIS A (sensory and motor complete) to at least AIS C (sensory and motor incomplete), the initial neurological evaluation had to be unambiguous to permit a posterior conclusion upon the efficacy of the treatment. To address this requirement, the initial diagnosis was carried out in two rounds, a preliminary and a confirmatory examination.

The preliminary examination was performed at the emergency setting by the on-call physician. It assessed only the assertive parameters of the AIS examination to define complete lesions, namely voluntary anal contraction (VAC), sensitivity to deep anal pressure (DAP) and light touch (LT) and pin prick (PP) sensitivities at sacral segments (S4-5). This is because evaluation of these four parameters is sufficient to identify a complete injury (AIS A). The confirmatory evaluation was done in the same manner (testing the four assertive criteria only) immediately before injection of the treatment. This second examination was performed by a neurosurgeon or physiatrist of the study team. The goal of the second confirmatory examination was to extend to maximal the time elapsed between the moment of the trauma and that of AIS ascription. This was, to the best of our understanding, the most appropriate way to conciliate the scientific demand of having a precise and unequivocal diagnosis (obtained 72h after trauma) [46] and the ethical imperative of not delaying the surgical treatment needed by the patient. By doing so, we assumed the risk of ending up with doubts about the initial diagnosis of the participant. For instance, if the patient were classified initially as AIS A, received the treatment 24 hours after trauma and changed to AIS B at the follow up of 48 hours, we would not be able to ascribe AIS progression to an effect of the treatment.

Evaluations done at 24 hours after trauma are not considered unambiguous [46], meaning that this hypothetical participant could in principle bear an actual incomplete, AIS B, injury from the beginning. This risk was considered unavoidable in the present study. In reality, we had two cases, in which the initial diagnosis was uncertain, exactly participants 1 and 3, who died during hospitalization. In all other cases, there was no doubt about the initial AIS A diagnosis. Participants 4, 6 and 7 received initial evaluation after 72 hours and participants 2, 5 and 8 remained AIS A at the 48 hours evaluation.

Follow-up examinations were performed by one trained physiatrist of the team and independently confirmed by a second one every time that there was doubt about the parameters collected. We found no divergences between the two examiners.

Patient care during hospitalization followed the standard procedures of each hospital, except for the additional examinations and data collection of the trial. None of the participants received methylprednisolone. Surgical decompression of the spinal cord and vertebral stabilization were done according to the patient’s need.

### Study medication

Intraparenchymal injections of polylaminin were carried out either during decompression surgery or percutaneously under X-ray guidance. Polylaminin at a final concentration of 100 μg/ml was prepared extemporaneously at the surgery site by mixing laminin (Merck-Millipore; catalogue number no. CC085) with vehicle (20 mM acetate buffer containing 1 mM CaCl_2_). The mixture was poured into a sterile large-mouth vessel and collected into a 1 mL syringe with a 26G needle. Polylaminin was delivered into the spinal cord, through two intramedullary injections, each containing half of the total volume of the dose. Injections were done in areas of preserved tissue rostral and caudal to the lesion epicentre. The injection depth and angle were defined by the surgeon as to make the two sites of fluid release as close as possible of the lesion site, previously determined by evaluation of MRI and/or CT images. Likewise, when there was no spinal cord exposure (patients that did not need surgery), two injections were performed, proximal and distally from the injury site, with the positioning of the needles guided by a C-arm x-ray intensifier.

A human equivalent dose (HED) [47] of 0.71 μg/kg was calculated based on the effective animal dose of 5 μg/kg, used in female Wistar rats (weighted 200-250g) and considering 70 kg as the average weight for humans.

HED = Animal dose (mg/kg) x (animal weight/human weight)^0.33^

This value was non-conservatively rounded up to 1 μg/kg based on the concept of micro dose for exploratory Phase 0 trials [48]. According to this concept, a single dose of up to 100 μg per individual (micro dose) can be used in Phase 0 studies, *i.e.*, in first-in-human trials of new investigational agents designed to obtain preliminary data mainly on drug targeting [49].

### Statistical analysis

Considering that this was a single-arm trial, performed to evaluate safety and efficacy in comparison to a universal baseline, the number of participants was defined as a convenience sample. The number of participants responded to budget limitations.

## Supporting information

Electromyography

## Data Availability

All data produced in the present study are available upon reasonable request to the authors

## ACKNOWLEDGEMENTS

We would like to thank the trial participants and their families. We are also grateful to Gabriella da Silva Branco, Marcelo da Silva Dehoul, Gabriel Soares Feydit Vieira, Marcelo Ares, Enio Kanayama, Gabriel Gomes Maia, Guilherme Galdino de Sousa, Raquel Marras, Catarina Freitas, Olavo Boher Amaral, and Martin Oudega for their help in data collection, patient care and/or scientific input We are also in debt with Fernanda Malheiros and Belmiro Tenório Siqueira for their dedicated assistance in animal studies. We acknowledge the support from Centro de Diagnóstico por Imagem (CDPI; Rio de Janeiro, Brazil) for providing MRI and CT exams at no cost and Allianz Seguros for the cost-free extension of the study’s insurance.

## AUTHORS CONTRIBUTIONS

Conceptualization and conduction of the animal study: ESL, OBF, MAN, CFS, LVA, RCLL, MAAMF, MR, AVG-S, TC-S

Conceptualization of the human study: KM, MABL, JRLM, AVG-S, BAC, PRL, TC-S

Conduction of surgeries and direction of patient care: MABL, BAC, AUCJ, PRL

Conduction of electrophysiological monitoring: ASF

Conduction of neurological evaluations: DRX, ACF

Conduction of physiotherapy: ADS

Data curation: KM, DRX, RMF, TC-S

Data analyses: KM, MABL, JRLM, GSH, ADS, MAN, ESL, ASF, TC-S

Funding acquisition: TC-S

Project administration: KM, RMF, TC-S

Writing: KM, JRLM, GSH, MAN, TC-S

## DATA AND MATERIALS AVAILABILITY

Relevant clinical information of the participants such as neurological records and original exams are available upon formal request from qualified scientific and medical researchers.

## LIST OF SUPPLEMENTARY MATERIALS

Supplementary Appendix

○ Supplementary Figures S1-S2
○ Supplementary Tables S1-S6

Supplementary Material 1: Electromyography exams

