## Supplemental Figures and Tables for "RETURN OF VOLUNTARY MOTOR CONTRACTION AFTER COMPLETE SPINAL CORD INJURY: A PILOT HUMAN STUDY ON POLYLAMININ"

Menezes et al., 2023

Supplementary Figures

2 Figures:
Figure S1 – Page 2-3
Figure S2 – Page 4

Supplementary Tables

6 Tables:
Table S1 – Page 5-6
Table S2 – Page 7-8
Table S3 – Pages 9
Table S4 – Pages 10-12
Table S5 – Page 13
Table S6 – Page 14

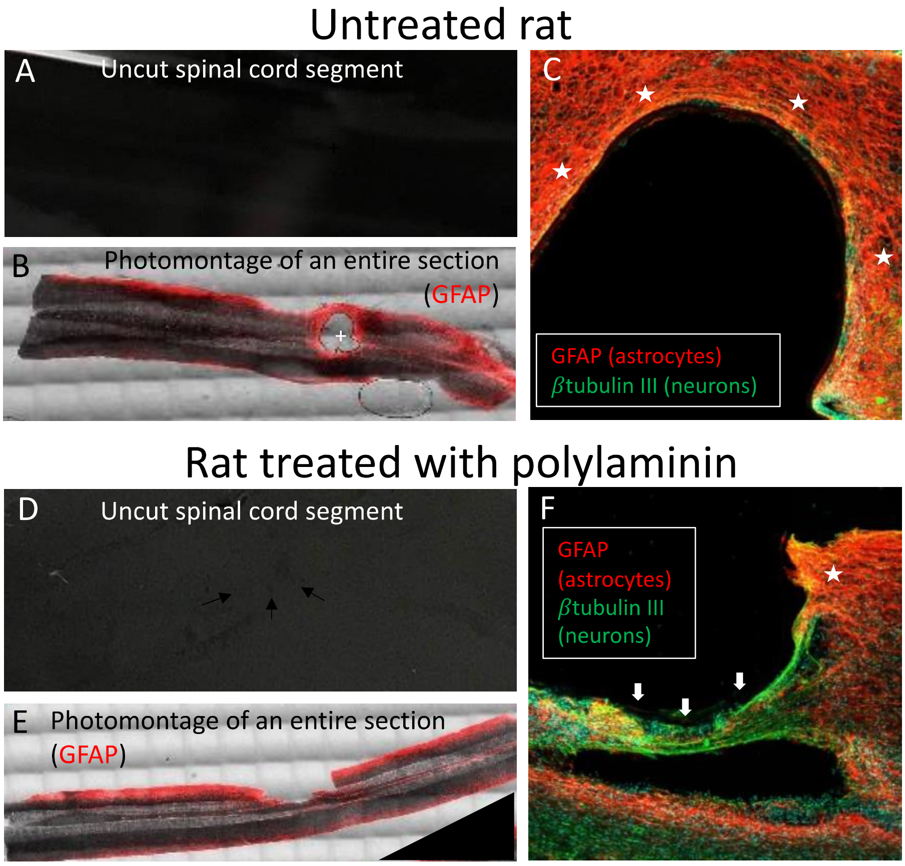

**Figure S1 – Morphological analysis of rat spinal cords.** The figure shows a comparison between two representative animals, not treated (A-C) and treated with polylaminin (D-F). After 8 weeks, rats were euthanized and had their vertebral columns removed, fixed and dissected to obtain the uncut segments of the spinal cords (A, D). The segments were sliced in a vibratome and 50 $\mu$m sections were immunostained to show areas occupied by reactive astrocytes (glial fibrillary acidic protein; GFAP), indicative of glial scaring (B, E). The control cord presents a rounded cavity, seen even in the uncut segment (crosses in panels A and B), surrounded by activated astrocytes (red in panel B). On the other hand, in the spinal cord of the animal receiving polylaminin, only a faint cavity-rim is visible in the uncut segment (arrows in panel D), while such cavity is not delineated by a strong reactivity for the astrocyte marker GFAP (red in panel E). Panels C and F show enlarged cavity areas (consecutive sections to B and E, respectively), where it can be appreciated that control animals display a strong and dense staining for GFAP (stars in panel C), whereas treated animals had a more confined expression of such marker (star in panel F). In addition, the polylamin-treated animal displays a tissue bridge across the lesion site (thick arrows in panel F). Regenerating axons (detected with the TUJ-1 antibody, more sensitive to $\beta$tubulin III in immature neurons; green) can be distinguished in association to the tissue bridge. Such bridge is not visible in the corresponding photo mounted image (E) probably because it does not expand throughout the whole thickness of the spinal cord and is visible only in fortuitous sections.

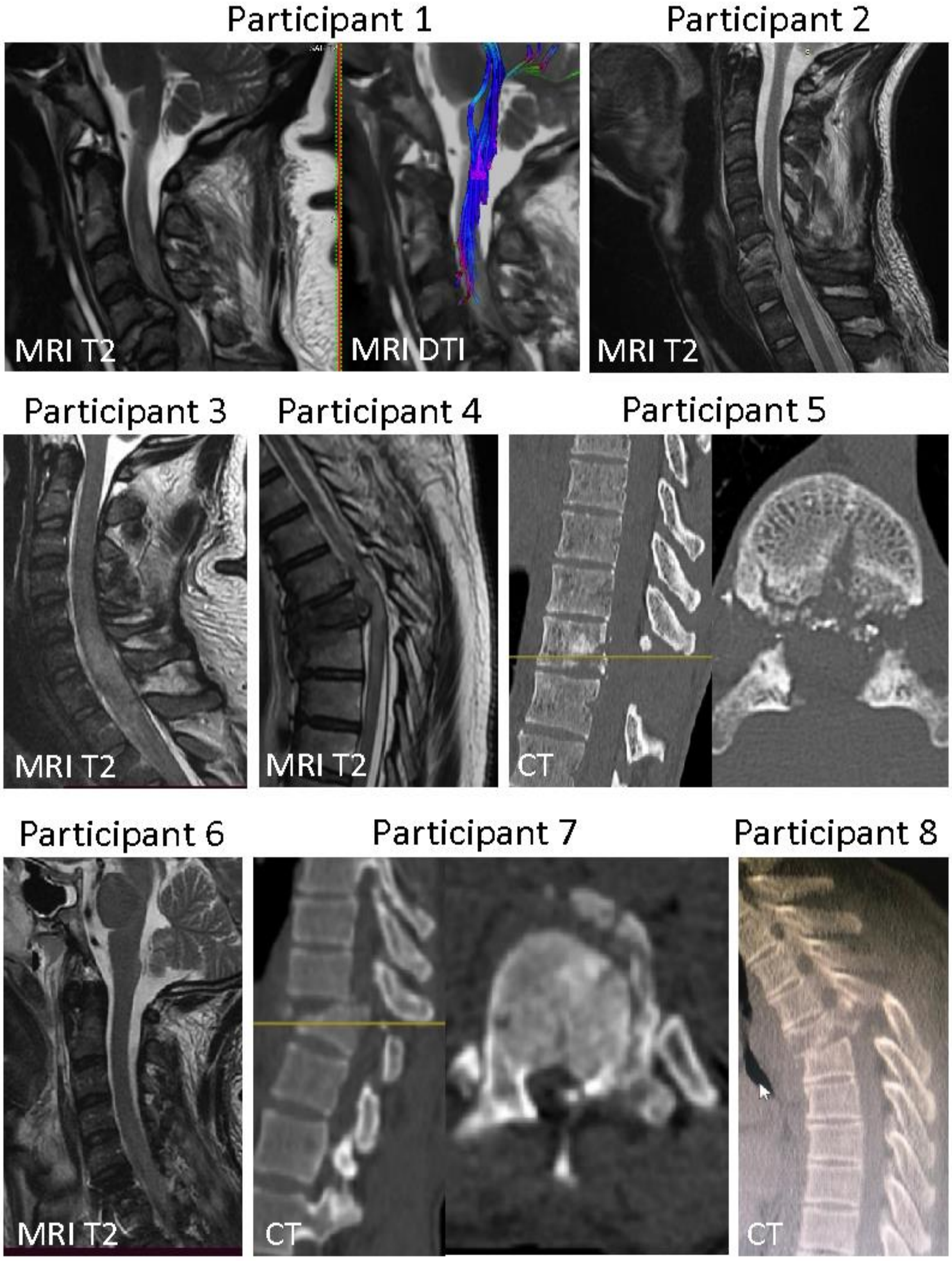

**Figure S2. Aspects of the initial lesions observed in MRI or CT.** Lesions of participants 1, 2, 3, 4 and 6 were evaluated by MRI. Lesions of participants 5, 7 and 8 were evaluated by CT due to the presence of projectile fragments (participant 5) or because the hospitalization centres had no MRI machine available (participants 7 and 8).

| *Complication type grouped by organ/system (N= 8)* | | *No. of complications* | *No. of patients* | *Incidence* |
|---|---|---|---|---|
| Infection | Pneumonia | 3 | 3 | |
| | Blood stream infection | 3 | 3 | |
| | Intestinal translocation infection | 1 | 1 | |
| | *Total of infections* | *7* | *5* | 63% |
| Cardiovascular | Arrhythmia<br>Bradycardia | 1<br>2 | 1<br>1 | |
| | Hypovolemic shock | 1 | 1 | |
| | Hypotension | 1 | 1 | |
| | Cardiorespiratory arrest | 2 | 2 | |
| | Cardiac tamponade | 1 | 1 | |
| | *Total of cardiovascular complications* | *8* | *2* | 25% |
| Pulmonary | Respiratory failure<br>Lobar collapse/atelectasis | 2<br>1 | 1<br>1 | |
| | Haemothorax | 2 | 2 | |
| | Hypoxemia | 1 | 1 | |
| | *Total of pulmonary complications* | *6* | *3* | 38% |
| Gastrointestinal | Rhabdomyolysis | 3 | 3 | |

| | | | | |
|---|---|---|---|---|
| Genitourinary (GI/GU) | Intestinal pseudo-obstruction | 1 | 1 | |
| | *Total of GI/GU complications* | *4* | *3* | 38% |
| Neurological/ Psychiatric (N/P) | Delirium and agitation | 2 | 2 | |
| | Reduced level of consciousness | 1 | 1 | |
| | *Total of N/P complications* | *3* | *2* | 25% |
| Haematological | Anaemia | 2 | 2 | |
| | *Total of haematological complications* | *2* | *2* | 25% |
| Skin | Pressure sores | 1 | 1 | |
| | *Total of skin complications* | *1* | *1* | 17% |

**Table S1. Severe complications in 8 patients (30 days after SCI).**

| Medical complications | | Classification of complication | N. of complications |
|---|---|---|---|
| **Infection** | Urinary tract infection | M (10[2,4,5,6,7,8]) | 10 |
| | Pyelonephritis | S (1[4]) | 1 |
| | Pneumonia | S (3[1,6,8]), M (1[3]) | 4 |
| | Genital Herpes infection | Mi (1[2]) | 1 |
| | Blood stream infection | S (3[2,5,8]), M(1[3]) | 4 |
| | Fungal skin infection | Mi (1[2]) | 1 |
| | Intestinal translocation infection | S (1[8]) | 1 |
| | Sepsis | S (1 [8]) | 1 |
| | **TOTAL** | | **23** |
| **Pulmonary** | Respiratory failure | S (2[1]) | 2 |
| | Lobar collapse/atelectasis | S (1[1]), M (1[4]) | 2 |
| | Haemothorax | S (2[4, 5]) | 2 |
| | Hypoxemia | S (1[1]) | 1 |
| | Pleural effusion | M (1[5]) | 1 |
| | Desaturation | M (1[5]) | 1 |
| | **TOTAL** | | **9** |
| **Neurological/ psychiatric** | Pain | Mi (2[1,5]), M(3[2,3,7]), | 5 |
| | Delirium and agitation | S (2[1,6]), M (2[2,8]) | 4 |
| | Headache | Mi (1[2]) | 1 |
| | Reduced level of consciousness | S (1[1]) | 1 |
| | Mental confusion | M (1[4]) | 1 |
| | Dysautonomia | M (1[2]) | 1 |
| | Vertigo | Mi (2[2,7]) | 2 |
| | Muscle spasm | M (1[2]), Mi (1[7]) | 2 |
| | Diabetes insipidus (polyuria) | M (1[6]) | 1 |
| | Drooling | Mi (1[6]) | 1 |
| | **TOTAL** | | **19** |
| **Haematological** | Anaemia | S (2[6,8]) | 2 |
| | **TOTAL** | | **2** |
| **Gastrointestinal and Genitourinary** | Nausea/vomiting | M (1[6]) | 1 |
| | Gastroparesis | M (1[6]) | 1 |
| | Rhabdomyolysis | S (3[2,6,8]) | 3 |
| | Constipation | Mi (5[2,4,5,7]) | 5 |
| | Intestinal pseudo-obstruction | S(1[8]) | 1 |
| | Heartburn | Mi (1[2]) | 1 |
| | Urinary retention | Mi (1[2]), M(1[4]) | 2 |
| | Pre-renal failure | M (1[3]) | 1 |
| | Anuria | M (1[3]) | 1 |

| | | | |
|---|---|---|---|
| | Haematuria | M ($1^{5}$) | 1 |
| | Diarrhoea | Mi ($1^{6}$) | 1 |
| | **TOTAL** | | **18** |
| **Skin** | Pressure-damage skin | M ($3^{2,7,8}$), S ($1^{4}$) | 4 |
| | Pruritus (pharmacology allergy) | Mi ($1^{4}$) | 1 |
| | Surgical wound hyperaemia | Mi ($1^{5}$) | 1 |
| | **TOTAL** | | **6** |
| **Cardiovascular** | Arrythmia | S ($1^{1}$) | 1 |
| | Bradycardia | S ($2^{1}$) | 2 |
| | hypovolemic shock | S ($1^{1}$) | 1 |
| | Septic shock | S ($1^{8}$) | 1 |
| | Hypotension | Mi($1^{1}$), S ($1^{1}$) | 2 |
| | Hypertension | M ($1^{8}$) | 1 |
| | Cardiorespiratory arrest | S ($3^{1,3,8}$) | 3 |
| | Cardiac tamponade | S ($1^{3}$) | 1 |
| | **TOTAL** | | **12** |
| **OTHERS** | Arthrodesis equipment poorly adapted by cervical tomography | Mi ($1^{1}$) | 1 |
| | Lower limb oedema associated with heterotopic ossification | M ($1^{5}$) | 1 |
| | lower limb oedema | M ($1^{8}$) | 1 |
| | **TOTAL** | | **3** |

**Table S2. All medical complications.** Medical complications observed in the 8 research participants over 12 months of clinical follow-up. The first column describes the types of complications, grouped by organ/system. The second column describes the classification of the complication as mild (Mi), moderate (M) or severe (S). The numbers in parentheses mean the number of events within each classification, and the superscripts identify the participants displaying the event. The third column describes the total number of complications in each category.

| | *Creatinine* *N (%)* | *Urea* *N (%)* | *ALT* *N (%)* | *AST* *N (%)* |
|---|---|---|---|---|
| **Before treatment** | | | | |
| Normal[1] | 7 (88) | 8 (100) | 3 (75) | 1 (20) |
| Mild[2] | 1 (12) | 0 (0) | 0 (0) | 3 (60) |
| Moderate[3] | 0 (0) | 0 (0) | 1 (25) | 1 (20) |
| Severe[4] | 0 (0) | 0 (0) | 0 (0) | 0 (0) |
| Total | 8 (100) | 8 (100) | 4 (100) | 5 (100) |
| **Two days after treatment** | | | | |
| Normal | 7 (88) | 7 (88) | 4 (80) | 1 (20) |
| Mild | 1 (12) | 1 (12) | 1 (20) | 3 (60) |
| Moderate | 0 (0) | 0 (0) | 0 (0) | 1 (20) |
| Severe | 0 (0) | 0 (0) | 0 (0) | 0 (0) |
| Total | 8 (100) | 8 (100) | 5 (100) | 5 (100) |
| **Ten days after treatment** | | | | |
| Normal | 7 (88) | 8 (100) | 2 (60) | 1 (20) |
| Mild | 1 (12) | 0 (0) | 3 (40) | 4 (80) |
| Moderate | 0 (0) | 0 (0) | 0 (0) | 0 (0) |
| Severe | 0 (0) | 0 (0) | 0 (0) | 0 (0) |
| Total | 8 (100) | 8 (100) | 5 (100) | 5 (100) |

**TABLE S3. Renal and hepatic markers before and after polylaminin administration.** [1]Normal applies to the following reference values: creatinine, 0.5-1.2 mg/dL; urea, 18-55 mg/dL; alanine aminotransferase (ALT), lower than 41 U/dL, and aspartate aminotransferase (AST), lower than 35 U/mL; [2]Mild: > upper limit of normal (ULN) to 2.5 x ULN; [3]Moderate: > 2.5-5 x ULN; [4]Severe: > 5-20 x ULN.

| Participant | | Initial (before treatment) | 1 day | 2 days | 1 month | 3 months | 6 months | 12 months | Reference values |
|---|---|---|---|---|---|---|---|---|---|
| 1 | Red cells (x10⁶/mm³) | 4.29 | 3.5 | 3.86 | | | | | 3.9 - 5.1 |
| | Haemoglobin (g/dL) | 11.1 | 9.4 | 10.8 | | | | | 12.0 - 16 |
| | Haematocrit (%) | 34.6 | 28.2 | 30.8 | | | | | 35 - 45 % |
| | Leukocytes (/mm³) | 11,100 | 12,620 | 15,690 | | | | | 4,000 – 10,000 |
| | Basophils (%) | 0.1 | 0 | 0 | | | | | 0 - 2 % |
| | Eosinophils (%) | 0.7 | 0 | 0 | | | | | 1 - 6 % |
| | Neutrophils (%) | 84.8 | 80.0 | 55 | | | | | 40 - 80 % |
| | Rods (%) | - | 5 | 1 | | | | | 0 - 5 % |
| | Segmented (%) | - | 75 | 54 | | | | | 40 - 80 % |
| | Lymphocytes (%) | 11.2 | 17 | 40 | | | | | 20 - 50 % |
| | Monocytes (%) | 3.2 | 3 | 5 | | | | | 2 - 10 % |
| | Platelets (/mm³) | 315,000 | 274,000 | 283,000 | | | | | 150,000 – 450,000 |
| 2 | Red cells (x10⁶/mm³) | 4.8 | 3.53 | 3.27 | 4.33 | 4.98 | 5.13 | 5.09 | 4.3 - 5.5 |
| | Haemoglobin (g/dL) | 15.1 | 11.1 | 10.6 | 13.9 | 14.8 | 16.2 | 16.1 | 13.5 - 18.0 |
| | Haematocrit (%) | 47.3 | 34.8 | 32.5 | 41.9 | 43 | 44.8 | 46.5 | 40 - 50% |
| | Leukocytes (/mm³) | 11,200 | 12,100 | 7,600 | 7,400 | 5,060 | 8,480 | 4,540 | 4,500 – 13,000 |
| | Basophils (%) | 0 | 0 | 0 | 0 | 1 | 0.2 | 0.4 | 0 - 2 % |
| | Eosinophils (%) | 1 | 0 | 2 | 2 | 4 | 0.4 | 2 | 1 - 6 % |
| | Neutrophils (%) | 90 | 83 | 69 | 75 | 55 | 79.9 | 49.3 | 40 - 80 % |
| | Rods (%) | 2 | 2 | 2 | 1 | 0 | - | 0 | 0 - 5 % |
| | Segmented (%) | 88 | 81 | 67 | 74 | 55 | - | 49.3 | 40 - 80 % |
| | Lymphocytes (%) | 6 | 12 | 24 | 15 | 34 | 9.4 | 41.9 | 20 - 50 % |
| | Monocytes (%) | 3 | 5 | 5 | 8 | 6 | 10.1 | 6.4 | 2 - 10 % |
| | Platelets (/mm³) | 167,000 | 133,000 | 129,000 | 126,000 | 128,000 | 145,000 | 118,000 | 150,000 – 450,000 |
| 3 | Red cells (x10⁶/mm³) | 3.89 | 3.47 | 3.55 | - | - | - | - | 4.3 - 5.5 |
| | Haemoglobin (g/dL) | 12.5 | 11 | 11.3 | - | - | - | - | 13.5 - 18.0 |
| | Haematocrit (%) | 36.3 | 31.8 | 32.3 | - | - | - | - | 40 - 50% |
| | Leukocytes (/mm³) | 8,300 | 6,500 | 8,000 | - | - | - | - | 4,500 – 13,000 |
| | Basophils (%) | 0 | 0 | 0 | - | - | - | - | 0 - 2 % |
| | Eosinophils (%) | 0 | 0 | 2 | - | - | - | - | 1 - 6 % |
| | Neutrophils (%) | 86 | 68 | 65 | - | - | - | - | 40 - 80 % |
| | Rods (%) | 2 | 1 | 1 | - | - | - | - | 0 - 5 % |
| | Segmented (%) | 86 | 67 | 64 | - | - | - | - | 40 - 80 % |
| | Lymphocytes (%) | 8 | 27 | 29 | - | - | - | - | 20 - 50 % |

| | | | | | | | | | |
|---|---|---|---|---|---|---|---|---|---|
| | Monocytes (%) | 4 | 5 | 4 | - | - | - | - | 2 - 10 % |
| | Platelets (/mm³) | 191,000 | 131,000 | 146,000 | - | - | - | - | 150,000 – 450,000 |
| 4 | Red cells (x10⁶/mm³) | 3.27 | 3.1 | 2.93 | 3.15 | 4.22 | 4.73 | 5.04 | 3.9 - 5.1 |
| | Haemoglobin (g/dL) | 9.6 | 8.7 | 8.5 | 8.2 | 10.5 | 12.4 | 13.5 | 12.0 - 16 |
| | Haematocrit (%) | 26.5 | 25.9 | 24.4 | 25.2 | 32 | 38.4 | 43.7 | 35 - 45 % |
| | Leukocytes (/mm³) | 7,200 | 8,400 | 7,600 | 11,900 | 4,800 | 6,900 | 5,120 | 4,000 – 10,000 |
| | Basophils (%) | 0.1 | 0 | 0 | 0 | 0 | 0 | 1 | 0 - 2 % |
| | Eosinophils (%) | 1 | 0 | 2 | 5 | 5 | 2 | 2 | 1 - 6 % |
| | Neutrophils (%) | 78.2 | 84 | 74 | 76 | 46 | 60 | 55 | 40 - 80 % |
| | Rods (%) | 2 | 6 | 6 | 2 | 3 | 3 | 2 | 0 - 5 % |
| | Segmented (%) | 75 | 78 | 68 | 74 | 43 | 57 | 53 | 40 - 80 % |
| | Lymphocytes (%) | 15 | 10 | 20 | 13 | 36 | 27 | 36 | 20 - 50 % |
| | Monocytes (%) | 7 | 6 | 4 | 6 | 13 | 11 | 6 | 2 - 10 % |
| | Platelets (/mm³) | 123,000 | 83,000 | 124,000 | 312,000 | 229,000 | 210,000 | 222,000 | 150.000 - 450.000 |
| 5 | Red cells (x10⁶/mm³) | 3.3 | 2.63 | 2.36 | 4.04 | - | 4.8 | 4.56 | 4.3 - 5.5 |
| | Haemoglobin (g/dL) | 9.7 | 7.7 | 7.5 | 11.6 | - | 13.4 | 13.2 | 13.5 - 18.0 |
| | Haematocrit (%) | 29.2 | 23.6 | 20.9 | 35 | - | 42.7 | 39 | 40 - 50% |
| | Leukocytes (/mm³) | 10,800 | 8,400 | 7,200 | 6,800 | - | 3,700 | 10,800 | 4,500 – 13,000 |
| | Basophils (%) | 0 | 0 | 0 | 0 | - | 0 | 0 | 0 - 2 % |
| | Eosinophils (%) | 0 | 2 | 1 | 2 | - | 2 | 3 | 1 - 6 % |
| | Neutrophils (%) | 90 | 73 | 75 | 64 | - | 46 | 65 | 40 - 80 % |
| | Rods (%) | 1 | 3 | 2 | 3 | - | - | 2 | 0 - 5 % |
| | Segmented (%) | 89 | 70 | 73 | 61 | - | - | 63 | 40 - 80 % |
| | Lymphocytes (%) | 3.1 | 20 | 18 | 29 | - | 47 | 25 | 20 - 50 % |
| | Monocytes (%) | 6.9 | 5 | 6 | 5 | - | 5 | 7 | 2 - 10 % |
| | Platelets (/mm³) | 162,000 | 115,000 | 123,000 | 447,000 | - | 307,000 | 315,000 | 150,000 – 450,000 |
| 6 | Red cells (x10⁶/mm³) | 2.73 | 2.91 | 3.4 | 3.73 | 3.66 | 4.22 | 4.59 | 4.3 - 5.5 |
| | Haemoglobin (g/dL) | 7.6 | 8.2 | 9.4 | 10.4 | 10.2 | 12.19 | 13.1 | 13.5 - 18.0 |
| | Haematocrit (%) | 24 | 25.9 | 29.5 | 33.1 | 31.6 | 39.5 | 40.4 | 40 - 50% |
| | Leukocytes (/mm³) | 5,000 | 7,020 | 5,610 | 7,020 | 4,630 | 4,770 | 4,720 | 4,500 – 13,000 |
| | Basophils (%) | 0 | 0 | 0 | 0 | 0.5 | 0.2 | 0.2 | 0 - 2 % |
| | Eosinophils (%) | 0 | 1 | 4.6 | 2 | 8.4 | 3.8 | 4.7 | 1 - 6 % |
| | Neutrophils (%) | 89 | - | - | - | 43.5 | - | - | 40 - 80 % |
| | Rods (%) | 5 | - | - | - | - | - | - | 0 - 5 % |
| | Segmented (%) | 84 | 83.2 | 66.7 | 60 | - | 54.9 | 53.4 | 40 - 80 % |
| | Lymphocytes (%) | 9 | 11.1 | 20.7 | 26 | 38.9 | 32.7 | 31.8 | 20 - 50 % |

| | | | | | | | | |
|---|---|---|---|---|---|---|---|---|
| | Monocytes (%) | 2 | 4.7 | 8 | 12 | 8.7 | 8.4 | 10 | 2 - 10 % |
| | Platelets (/mm$^3$) | 123,000 | 128,000 | 155,000 | 248,000 | 218,000 | 206,000 | 193,000 | 150,000 – 450,000 |
| 7 | Red cells (x10$^6$/mm$^3$) | 3.6 | 2.9 | 2.9 | 3.21 | 4.7 | 4.14 | 4.34 | 3.9 - 5.1 |
| | Haemoglobin (g/dL) | 11 | 8.8 | 8.9 | 11.3 | 12.8 | 12 | 13.4 | 12.0 - 16 |
| | Haematocrit (%) | 33.8 | 28.7 | 28.8 | 32.7 | 40.6 | 35.7 | 39.6 | 35 - 45 % |
| | Leukocytes (/mm$^3$) | 8,500 | 10,500 | 9,300 | 10,750 | 10,000 | 13,180 | 9,370 | 4,000 – 10,000 |
| | Basophils (%) | 0 | 0 | 0 | 0 | 0 | 0 | 0.2 | 0 - 2 % |
| | Eosinophils (%) | 0 | 0 | 0 | 3 | 8 | 1 | 1.3 | 1 - 6 % |
| | Neutrophils (%) | 82 | 70 | 97 | 65 | 57 | 85 | 64.4 | 40 - 80 % |
| | Rods (%) | 3 | 3 | 27 | 2 | 0 | 3 | - | 0 - 5 % |
| | Segmented (%) | 79 | 67 | 70 | 65 | 57 | 82 | - | 40 - 80 % |
| | Lymphocytes (%) | 12 | 18 | 2 | 23 | 28 | 12 | 26.7 | 20 - 50 % |
| | Monocytes (%) | 6 | 12 | 10.1 | 7 | 7 | 2 | 7.4 | 2 - 10 % |
| | Platelets (/mm$^3$) | 244,000 | 266,000 | 227,000 | 349,000 | 330,000 | 316,000 | 259,000 | 150,000 – 450,000 |
| 8 | Red cells (x10$^6$/mm$^3$) | 4.31 | - | 3.37 | 3.19 | | | | 4.3 - 5.5 |
| | Haemoglobin (g/dL) | 13.3 | - | 10.6 | 9.7 | | | | 13.5 - 18.0 |
| | Haematocrit (%) | 38.8 | - | 30.4 | 26.3 | | | | 40 - 50% |
| | Leukocytes (/mm$^3$) | 11,370 | - | 13,160 | 26,030 | | | | 4,500 – 13,000 |
| | Basophils (%) | 2 | - | 0 | 0 | | | | 0 - 2 % |
| | Eosinophils (%) | 0 | - | 2 | 0 | | | | 1 - 6 % |
| | Neutrophils (%) | 110 | - | 83 | 89 | | | | 40 - 80 % |
| | Rods (%) | 3 | - | 3 | 7 | | | | 0 - 5 % |
| | Segmented (%) | 80 | - | 80 | 82 | | | | 40 - 80 % |
| | Lymphocytes (%) | 11 | - | 10 | 8 | | | | 20 - 50 % |
| | Monocytes (%) | 4 | - | 5 | 3 | | | | 2 - 10 % |
| | Platelets (/mm$^3$) | 133,000 | - | 101,000 | 435,000 | | | | 150,000 – 450,000 |

**TABLE S4. Blood counts before and after polylaminin administration.**

| *Study sites* | *Included participants* |
|---|---|
| **Public Hospitals** | |
| Public State Hospital Azevedo Lima (HEAL) | 1, 8 |
| Army Central Hospital (HCE) | 3 |
| Rio de Janeiro Municipal Hospital Souza Aguiar (HMSA) | 4, 5[a], 7 |
| **Private Hospitals** | |
| Hospital Copa Star (HCS) | 2 |
| Hospital Ênio Serra (HES) | 5[b] |
| Hospital São José (HSJ) | 6[a] |
| Hospital Vila da Serra (HVS) | 6[b] |

**Table S5. Study sites**. [a] hospital of inclusion, [b] hospital of long-term hospitalization

*Inclusion criteria*

- Age between 18 and 70 years old
- ASIA Impairment Scale grade A
- Traumatic spinal cord injury at neurologic level C4 to T12
- Written informed consent by patient or legally authorized representative to participate in the study

*Exclusion criteria*

- Severe brain trauma
- Need of permanent mechanical respiratory support
- Concomitant trauma considered by the study physicians to be severe enough to preclude neurological improvement
- Previous neurological diseases (EX. Alzheimer's, Amyotrophic Lateral Sclerosis, Cerebrovascular Accident sequelae) or previous neurological deficits of any aetiology
- Abuse or dependence of alcohol or illicit drugs
- Any other comorbidity that, at the discretion of the investigator, precludes inclusion of the patient in the study

**Table S6. Eligibility criteria.**
