## Supplementary material for "RETURN OF VOLUNTARY MOTOR CONTRACTION AFTER COMPLETE SPINAL CORD INJURY: A PILOT HUMAN STUDY ON POLYLAMININ": Electromyography

### PARTICIPANT 2

#### INITIAL

| | Right side (RMS V) | Left side (RMS V) |
|---|---|---|
| Biceps brachii | 1.2967493 | NaN |
| Triceps brachii | 0.1589289 | NaN |
| Quadriceps femoris | NaN | NaN |
| Tibialis anterior | 0.2226789 | NaN |
| Gastrocnemius medial | 0.1954454 | NaN |

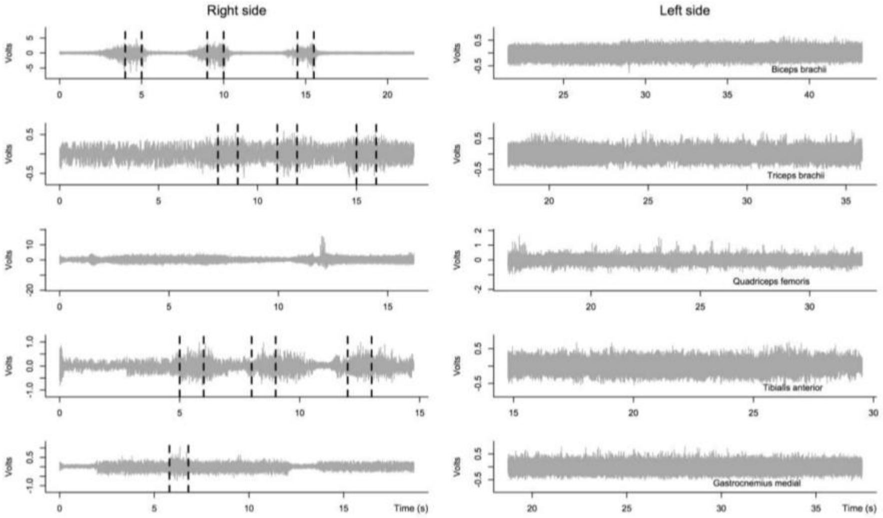


#### FINAL

| | Right side (RMS V) | Left side (RMS V) |
|---|---|---|
| Biceps brachii | 0.16511261 | 0.138843771 |
| Triceps brachii | 0.93459217 | 0.967289973 |
| Quadriceps femoris | 0.16513402 | 0.033278241 |
| Tibialis anterior | 0.02001822 | 0.006317445 |
| Gastrocnemius medial | 0.05740483 | 0.015812209 |

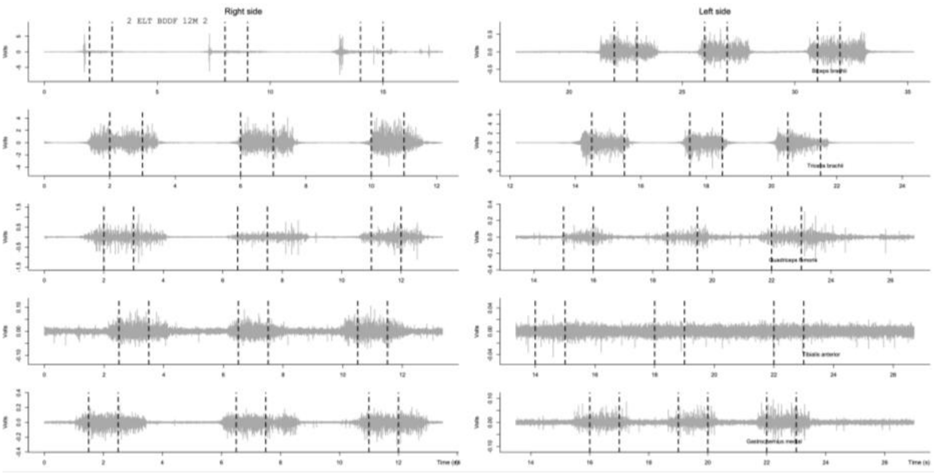

### PARTICIPANT 4

#### INITIAL

| | Right side (RMS V) | Left side (RMS V) |
|---|---|---|
| Biceps brachii | NaN | NaN |
| Triceps brachii | 0.2072178 | NaN |
| Quadriceps femoris | NaN | NaN |
| Tibialis anterior | NaN | NaN |
| Gastrocnemius medial | NaN | NaN |

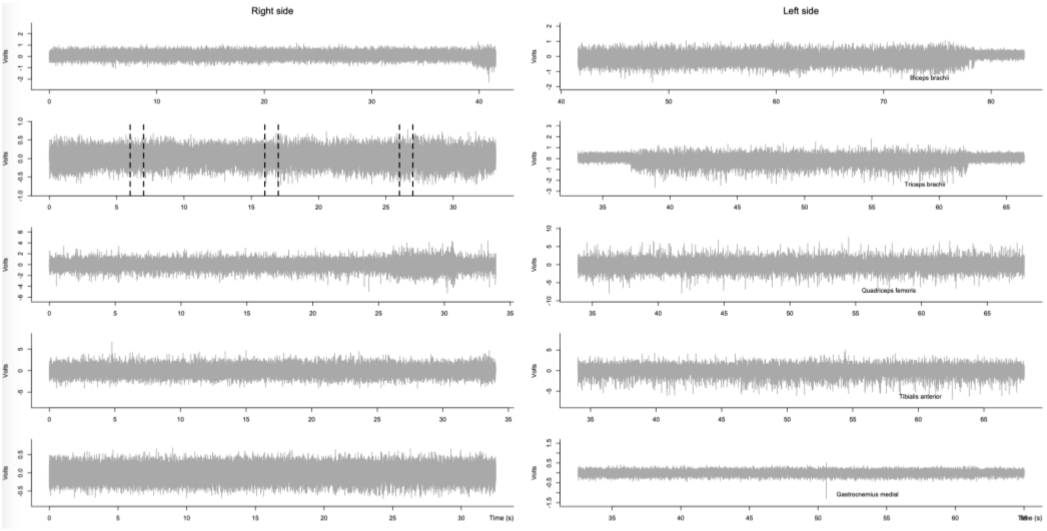


#### FINAL

| | Right side (RMS V) | Left side (RMS V) |
|---|---|---|
| Biceps brachii | 0.1532704 | 0.3214382 |
| Triceps brachii | 0.3773685 | 0.2684005 |
| Quadriceps femoris | NaN | NaN |
| Tibialis anterior | NaN | NaN |
| Gastrocnemius medial | NaN | NaN |

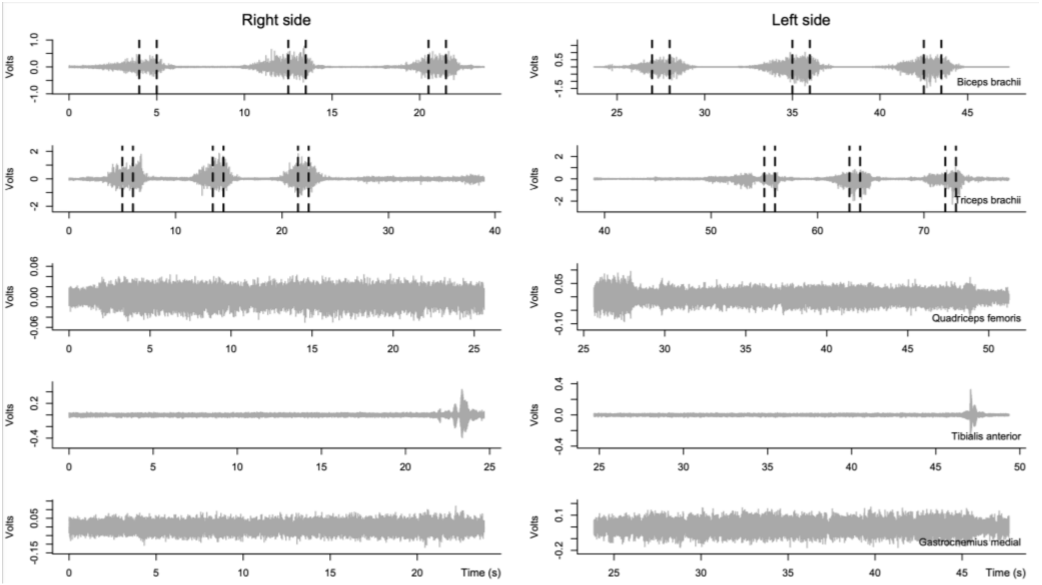

### PARTICIPANT 5

#### INITIAL

| | Right side (RMS V) | Left side (RMS V) |
|---|---|---|
| Biceps brachii | 1.337749 | 2.218984 |
| Triceps brachii | 1.795759 | 1.289479 |
| Quadriceps femoris | NaN | NaN |
| Tibialis anterior | NaN | NaN |
| Gastrocnemius medial | NaN | NaN |

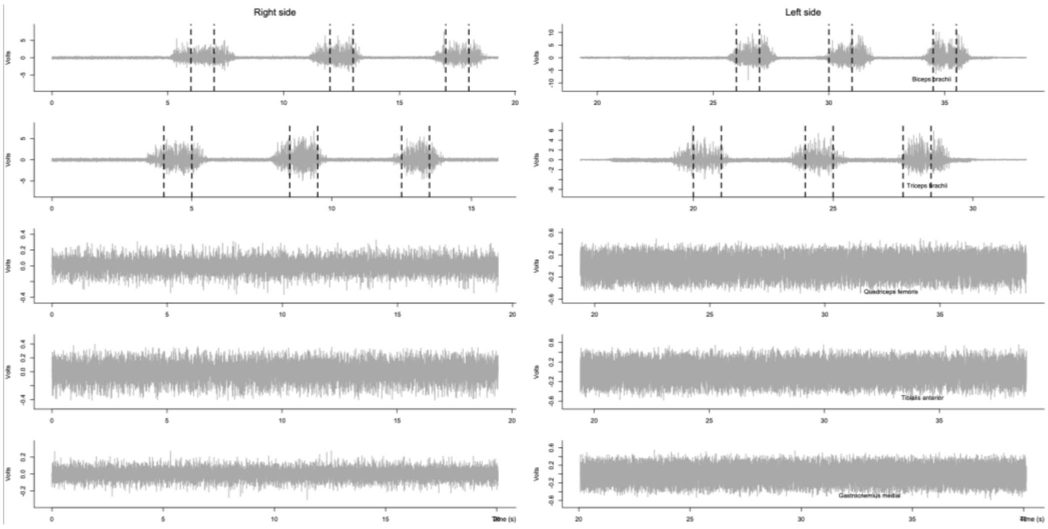


#### FINAL

| | Right side (RMS V) | Left side (RMS V) |
|---|---|---|
| Biceps brachii | 0.9227677 | 0.8632993 |
| Triceps brachii | 1.3548200 | 0.8365979 |
| Quadriceps femoris | NaN | NaN |
| Tibialis anterior | NaN | NaN |
| Gastrocnemius medial | NaN | NaN |

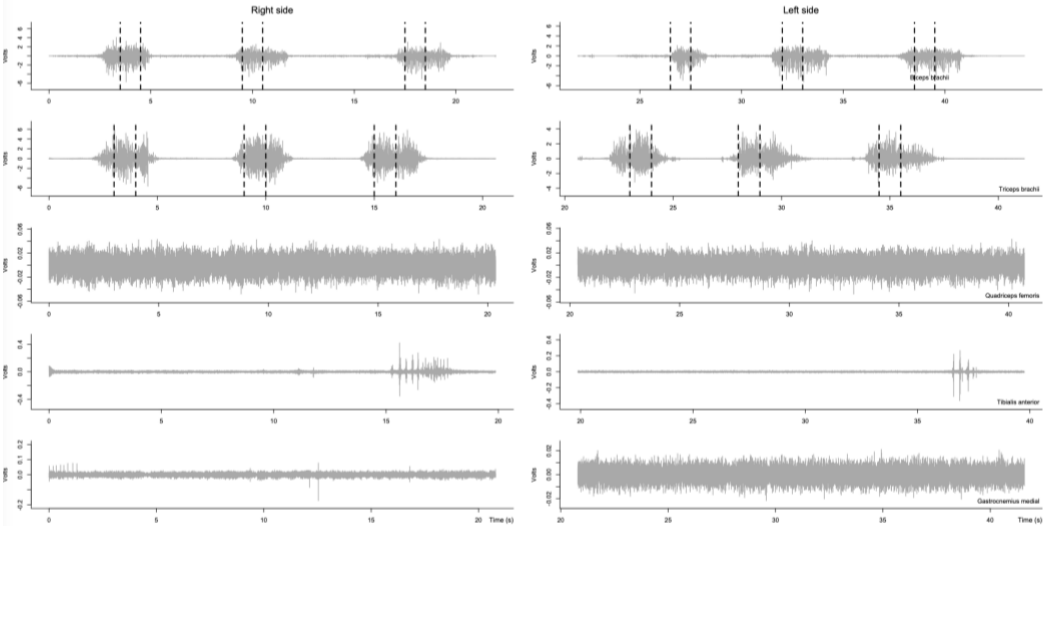
